# The origin, epidemiology and phylodynamics of HIV-1 CRF47_BF

**DOI:** 10.1101/2022.03.08.22272034

**Authors:** Gracelyn Hill, Marcos Pérez-Losada, Elena Delgado, Sonia Benito, Vanessa Montero, Horacio Gil, Mónica Sánchez, Javier Cañada-García, Elena García-Bodas, Keith A. Crandall, Michael M Thomson, The Spanish Group for the Study of New HIV Diagnoses

**Author notes:** **Correspondence:** Michael Thomson,; Keith Crandall. **Author Emails:** Gracelyn Hill; Marcos Pérez-Losada; Elena Delgado; Sonia Benito; Vanessa Montero; Horacio Gil; Mónica Sánchez; Javier Cañada; Elena García-Bodas; Keith Crandall; Michael Thomson.

## Abstract

CRF47_BF is a circulating recombinant form (CRF) of the human immunodeficiency virus type 1 (HIV-1), the etiological agent of AIDS. CRF47_BF represents one of 19 CRFx_BFs and has a geographic focus in Spain, where it was first identified in 2010. Since its discovery, CRF47_BF has expanded considerably in Spain, predominantly through heterosexual contact (∼56% of the infections). Little is known, however, about the origin and diversity of this CRF or its epidemiological correlates, as very few samples have been available so far. This study conducts a phylogenetic analysis with representatives of all CRFx_BF sequence types along with HIV-1 M Group subtypes to place the CRF47_BF sequences in a definitive phylogenetic context. The CRFx_BF sequences cluster into a single, not well supported, clade that includes their dominant parent subtypes (subtype B and subtype F). This clade also includes subtype D and excludes subsubtype F2. The CRF47_BF sequences all share a most recent common ancestor. Further analysis of this clade couples CRF47_BF protease-reverse transcriptase sequences and epidemiological data from an additional 87 samples collected throughout Spain, coupled with additional CRF47_BF database sequences from Brazil and Spain to investigate the origin and phylodynamics of CRF47_BF. The Spanish region with the highest proportion of CRF47_BF samples in the data set was the Basque Country (43.7%) with Navarre next highest at 19.5%. We include in our analysis epidemiological data on host sex, mode of transmission, time of collection, and geographic region. The phylodynamic analysis indicates that CRF47_BF originated in Brazil around 1993-1994 and spread to Spain from Brazil in approximately 1999-2000. The virus spread rapidly throughout Spain with increasing population sizes prior to 2010 and again between 2010 and 2017 with population declines to 2019 and a steady state through 2020. Three strongly supported clusters associated with Spanish regions (Basque Country, Navarre, and Aragon), together comprising 60.8% of the Spanish samples, were identified, one of which was also associated with transmission among men who have sex with men. The expansion in Spain of CRF47_BF, together with that of other CRFs and subtype variants of South American origin, previously reported, reflects the increasing relationship between the South American and European HIV-1 epidemics.

## 1 Introduction

High genetic diversity of HIV-1 is a defining feature of the AIDS virus. This diversity gain and loss is a hallmark of the evolution of HIV in the context of drug resistance and changing environments (Pennings et al., 2014). A contributing factor in the evolution of HIV is the process of recombination (Rambaut et al., 2004; Vuilleumier and Bonhoeffer, 2015). Genetic recombination is known to impact HIV allelic diversity and subsequent population dynamics at a rate equivalent to the high mutation rate of HIV (Shriner et al., 2004). Genetic diversity within HIV subtypes can be up to 17% sequence divergence across the genome with 17-35% divergence between subtypes (Castro-Nallar et al., 2012a). Yet recombination can even occur between subtypes as HIV variants spread around the globe, leading to circulating recombinant forms or CRFs, as well as unique recombinant forms (URFs) (Castro-Nallar et al., 2012b). There are currently 118 known HIV-1 CRFs according to the Los Alamos HIV Sequence Database (Los Alamos National Laboratory - HIV Databases) involving recombination events between nearly all known subtypes and even between other CRFs (e.g., CRF15_01B is a recombinant form between CRF01 and subtype B (Tovanabutra et al., 2003)). The CRFs often have their own unique population dynamics and molecular epidemiology compared to their parental strains and often lead to novel infection dynamics and spread. One such CRF is CRF47_BF, discovered in Spain and described in 2010 (Fernández-García et al., 2010) as an intersubtype recombinant form between HIV-1 subtypes B and F. Of the CRFs, among the most abundant are those between B and F subtypes, with 19 CRF_BFs (note that in the Los Alamos HIV Database these are sometimes designated ‘BF’ and sometimes ‘BF1’, even for the same CRF). Of the CRF_BFs, all but two are known from South America (mainly Brazil, but Argentina, Uruguay, Paraguay, Chile, Peru, and Bolivia as well) with a few found both in South America and Europe (CRF66, 75, and 89). Only two CRF_BF have been reported to be found circulating exclusively in Europe, CRF42_BF in Luxembourg (Struck et al., 2015) and CRF47_BF in Spain (Fernández-García et al., 2010). Since its description, CRF47_BF has expanded considerably in Spain, predominantly via heterosexual contact, and is now known from Brazil as well, as attested by a CRF47_BF virus collected in this country whose sequence is deposited in the Los Alamos database (Los Alamos National Laboratory - HIV Databases).

The goal of this study is to estimate the temporal and geographic origin of CRF47_BF and estimate the dynamics of diffusion and growth throughout its evolutionary history. Towards this goal, we combine new CRF47_BF sequence data from our lab from strains isolated in Spain with data from other BF strains in the Los Alamos database to place CRF47_BF in the context of the 19 CRF_BFs, and B, F, and other subtypes.

## 2 Methods

### 2.1 Sample and data collection

Plasma and whole blood samples were collected from HIV-1-infected patients at public hospitals across 8 regions in Spain. Epidemiological data from the CRF47_BF patients were collected to link to the HIV sequence data. The epidemiological data included patient gender, the transmission route, the patient’s year of HIV diagnosis and date of sample collection, the region from which the sample was collected, the country of origin of the individual, and whether the patient was on antiretroviral (ARV) therapy.

The study was approved by the Committee of Research Ethics of Instituto de Salud Carlos III, Majadahonda, Madrid, Spain (report numbers CEI PI 38_2016-v3 and CEI PI 31_2019-v5). The study did not require written informed consent by the study participants, as it used samples and data collected as part of routine clinical practice and patients’ data were anonymized without retaining data allowing individual identification.

### 2.2 Sequence Analysis

(RT-)PCR was used to amplify the protease-reverse transcriptase (PR-RT) gene region from plasma-extracted RNA or whole blood-extracted DNA using previously described primers (Delgado et al., 2015) (Figure 1). PCR products were sequenced using the Sanger method with an automated capillary sequencer. These data were combined with PR-RT sequences classified as CRF47_BF at the Los Alamos HIV Sequence Database and reference sequences for all subtypes and all CRFx_BFs for this same gene region from the Los Alamos HIV Database. Finally, we conducted a BLAST (Altschul et al., 1990) search against GenBank with the 5’-most 950 nt of PR-RT of CRF47_BF viruses and included all sequences within 95% similarity.

**Figure 1.**
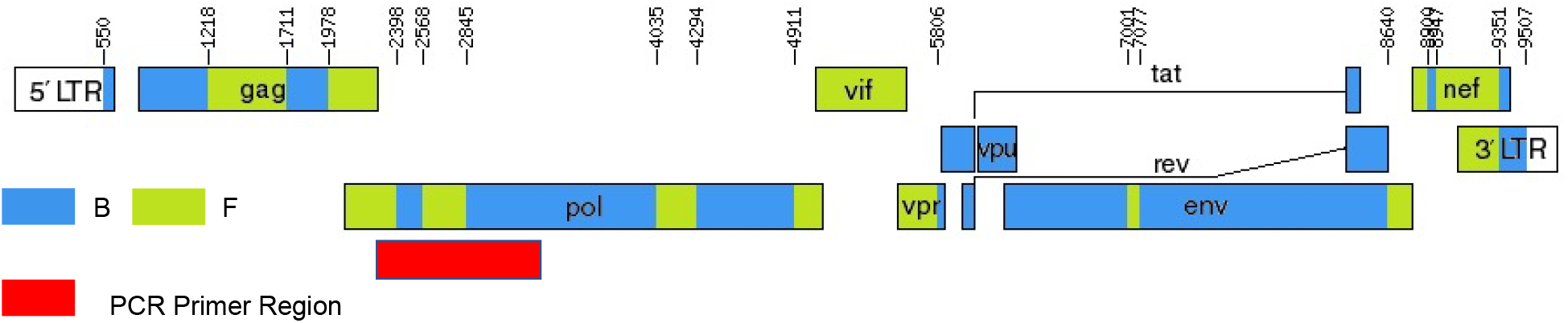
Mosaic genomic structure of CRF47_BF (from Los Alamos National Lab HIV Database) with targeted primers for the PR_RT amplicon annotated (shown in red) on the *pol* gene.

We conducted two analyses with these data. First, we included all data to validate the quality of the data and place the CRF47_BF within a broader phylogenetic context. Second, we conducted a focused analysis on the targeted CRF47_BF strains. In both analyses, we aligned sequence data using MAFFT (Katoh and Standley, 2013) with the FFT-NS-2 progressive alignment approach since these sequences are relatively similar. Phylogenetic analyses were conducted using maximum-likelihood (Felsenstein, 1981; Posada and Crandall, 2021) as implemented by RAxML (Kozlov et al., 2019) via the CIPRES web service (Miller et al., 2012). The phylogenetic analyses utilized the best-fit model of evolution (Posada and Crandall, 1998) as determined by ModelTest-NG (Darriba et al., 2020). Phylogenetic analyses were also done using a Bayesian approach as implemented by MrBayes 3.2 (Ronquist et al., 2012) with integrated model selection, 10 million MCMC generations, and codon partitioning. Confidence in the resulting phylogenetic estimates was assessed using the bootstrap approach (Felsenstein, 1985) for the maximum-likelihood analyses with 1,000 pseudoreplicates and with posterior probabilities (pP) in the Bayesian framework. Phylogenetic trees were visualized with iTOL (Letunic and Bork, 2019), as well as mapping of epidemiological characters along the phylogeny. For the focused CRF47_BF analyses, BEAST 2 (Bouckaert et al., 2014) was used to estimate a chronogram and the phylodynamic history of the CRF47_BF sequences with 10 million generations, codon partitioning, a strict clock model, estimated base frequencies and verification of convergence using Tracer (Rambaut et al., 2018). The input file used for this phylogenetic analysis was created using BEAUti. Codon partitioning for all analyses was divided such that the last nucleotide in the codon was a separate partition. Past population dynamics was estimated via Skygrid analysis (Hill and Baele, 2019), with BEAUti again being used for the creation of the input file. An HKY+G evolution model was used, along with codon partitioning, an uncorrelated relaxed clock model, and coalescent Bayesian Skygrid tree priors. The program Tracer was used to visualize the Skygrid plot after the analysis was completed. Finally, known drug resistant mutations were identified in the CRF47_BF data using the Stanford HIV Drug Resistance Database’s HIVdb v9.0 program (Tang et al., 2012).

Our initial phylogenetic analysis included subtypes from the HIV-1 M group (subtypes A1, A2, B, C, D, F1, F2, G, H, J, K, and L), as well as the CRF_BF recombinants. Our final alignment included 14 sequences representing all the major subtypes within HIV-1 group M, 5 subtype B sequences, 11 subtype F (F1, F2) sequences, and 34 representatives of all known and distinct CRF_BFs. In addition, we included all 98 sequences from CRF47_BF, including 87 obtained by us (7 from a previous study (Fernández-García et al., 2010) and 80 newly derived) from the patients summarized in Table 1, and 12 from databases (10 from Spain and 2 from Brazil).

**Table 1.** Summary data from patients with CRF47_BF variant from Spain.

|  | Total N=87 | Percent |
| --- | --- | --- |
| **Gender** |  |  |
| Male | 68 | 78.2 |
| Female | 18 | 20.7 |
| Transgender | 1 | 1.2 |
| **Region** |  |  |
| Basque Country | 38 | 43.7 |
| Navarre | 17 | 19.5 |
| Galicia | 14 | 16.1 |
| Aragon | 9 | 10.3 |
| Comunitat Valenciana | 6 | 6.9 |
| Castilla y Leon | 1 | 1.2 |
| Castilla-La Mancha | 1 | 1.2 |
| Madrid | 1 | 1.2 |
| **Transmission route** |  |  |
| Heterosexual | 49 | 56.3 |
| MSM | 19 | 21.8 |
| Sexual Transmission (Unspecified Sexuality) | 12 | 13.8 |
| Other/No Data | 7 | 8.1 |
| **ARV Therapy** |  |  |
| No | 75 | 86.2 |
| Yes | 7 | 8.1 |
| No Data | 5 | 5.7 |
| **Country of Origin** |  |  |
| Spain | 68 | 78.2 |
| Brazil | 5 | 5.8 |
| Colombia | 5 | 5.8 |
| Other | 4 | 4.6 |
| Morocco | 3 | 3.5 |
| Nicaragua | 2 | 2.3 |

### 2.3 Statistical analyses

Correlations between cluster membership and epidemiological data were analyzed with Fisher’s exact test.

## 3 Results

### 3.1 Epidemiology

We collected samples and epidemiological data from 87 patients throughout 8 different regions of Spain (Basque Country, Navarre, Galicia, Aragon, Comunitat Valenciana, Madrid, Castilla-La Mancha, and Castilla y León). Collections were made from 2010 to 2021. Males accounted for 78% of the individuals with CRF47_BF in our study and 56% of individuals reported transmission via heterosexual contact (61% considering only individuals with available data on transmission route) (Table 1). The Spanish region with the highest proportion of the CRF47_BF variant in our data set was the Basque Country with 44% of the cases, while Navarre was the next highest (18% of the cases). Most samples were collected shortly after HIV diagnosis. Patients received ARV therapy after sample collection.

### 3.2 Phylogenetics

The first phylogenetic analysis was a maximum likelihood phylogenetic estimate of the relationships amongst the CRFx_BFs, including HIV-1 M subtypes as outgroup taxa and subtypes B, F, and CRFx_BFs as ingroup taxa. Our RAxML tree depicted a monophyletic cluster of the subtype B, F, and CRF_BFs relative to the other HIV-1 subtypes (Figure 2), but including also Subtype D. The backbone structure of the CRF phylogenetic relationships was weakly supported (<70% bootstrap support – indicated by dashed lines), which is not particularly surprising given the potential difficulty in representing evolutionary histories of recombinant HIV-1 forms as bifurcating trees (Posada and Crandall, 2001, 2002). Many of the CRFx_BF forms cluster in strongly supported monophyletic groups themselves (e.g., CRF40_BF, CRF72_BF, CRF75_BF, CRF90_BF, CRF89_BF, etc.), including our target group of CRF47_BF sequences. Many of the other CRFs form weakly supported monophyletic groups (e.g., CRF70_BF, CRF46_BF, CRF38_BF, etc.) and a few form non-monophyletic groupings (e.g., CRF66_BF, CRF71_BF). The subtype B sequences cluster together within the CRFx_BF clade with both a cluster of subtype D and the CRF28_BF sequence nested within this subtype B cluster. Nevertheless, the target group for this study, the CRF47_BF sequences, clearly form a monophyletic group, suggesting independent evolution, and are a sister group to the CRF44_BF clade.

**Figure 2.**
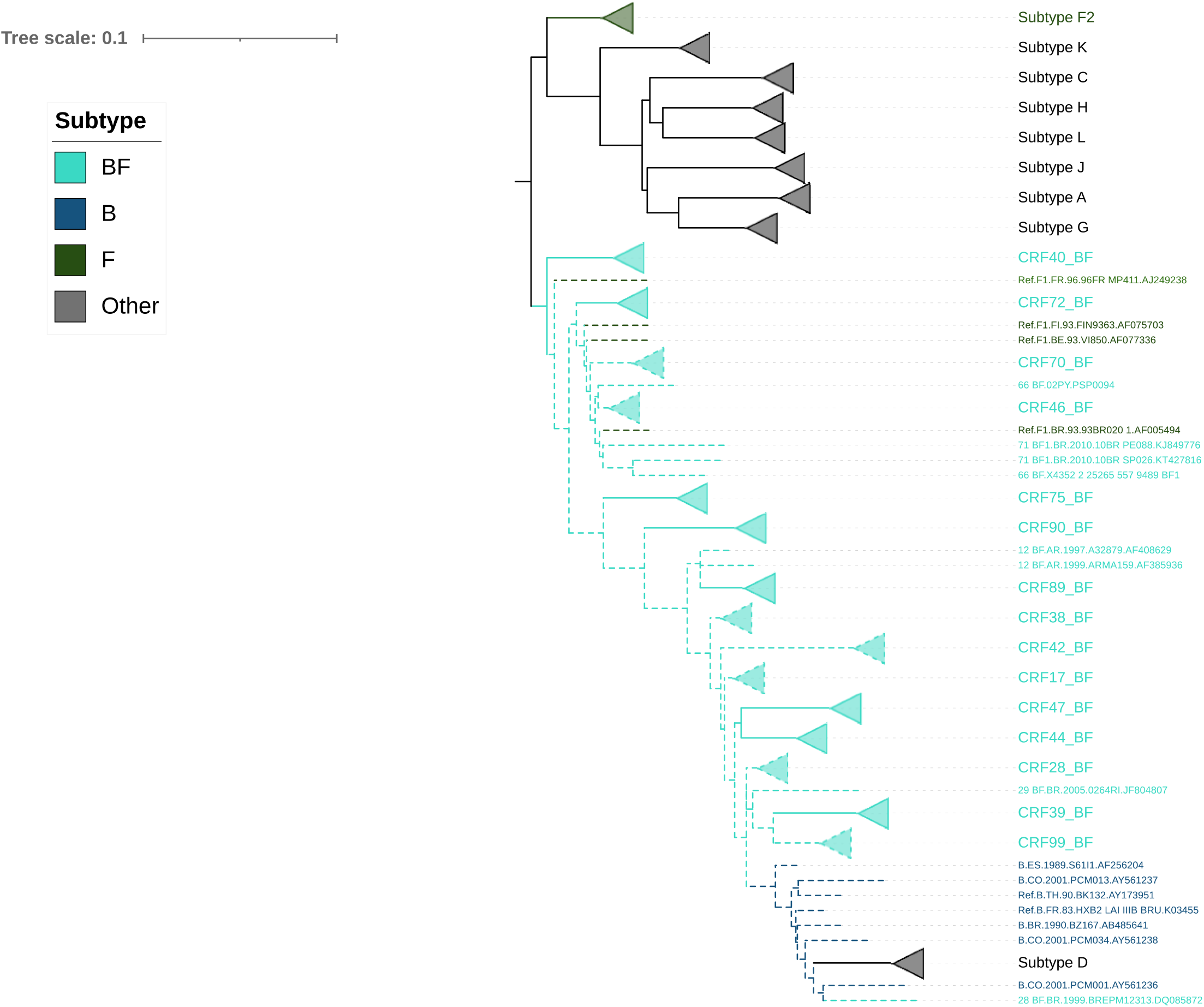
Maximum likelihood estimate of phylogenetic relationships amongst the HIV-1 M group subtypes and the CRFx_BFs with a focus on subtypes B and F. Lineages shown with dashed lines have <70% bootstrap support, whereas lineages shown in solid lines have ≥70% bootstrap support.

The Bayesian estimated phylogeny for the CRF47_BF sequences shows a monophyletic grouping of the sequences from Spain (Figure 3) with the two sequences from Brazil branching basally (KJ849798 and JQ238096). Within the Spanish cluster, there are three strongly supported clusters, comprising 29 (cluster I), 17 (cluster II), and 13 (cluster III) viruses, respectively, which are associated with the Basque Country (p=0.0002), Navarre (p=0.0001), and Aragon (p=0.0002), respectively. This is indicative of a single introduction of CRF47_BF into Spain with subsequent spread throughout the country and point introductions with subsequent expansion in different regions. The mixing of patient gender throughout the resulting phylogeny supports the epidemiological data suggesting predominantly heterosexual transmission among patients. We also found that cluster II, associated with Navarre, was associated with men who have sex with men (MSM) (p=0.0388). In this cluster, 14 of 15 individuals with known gender are men.

**Figure 3.**
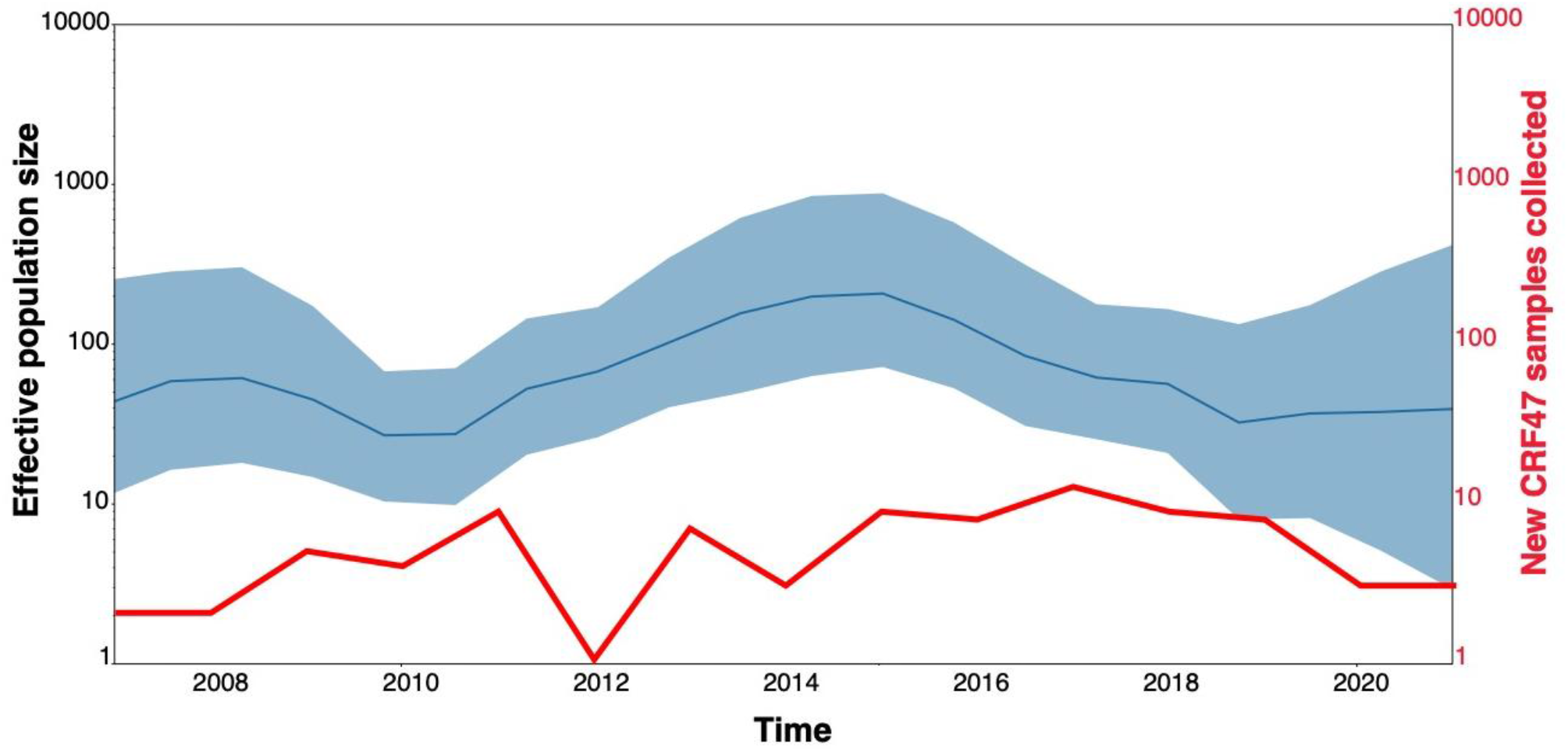
Bayesian (MrBayes) phylogenetic estimate of CRF47_BF sequences from Spain (colored by region) and other CRF47_BF sequences from GenBank, as well as a few additional Subtype B sequences from Spain, Brazil, and Colombia plus HXB2, all to serve as an outgroup to the CRF47 sequences. Only clade posterior probabilities <0.95 are indicated by an *; all other clades showed posterior probabilities ≥0.95. Epidemiological data are mapped to the right of the phylogeny, including days from diagnosis., Sex, Transmission, and Geographic Region. Branch lengths are drawn proportional to the amount of sequence divergence. Clusters corresponding to the Spanish regions of Basque Country (cluster I), Navarre (cluster II), and Aragon (cluster III) are indicated.

Based on the sample dates, we grouped these in four temporal categories of recency (days between diagnosis date and current date) (>3000, 2000-3000, 1000-2000, and <1000 days from current). Thus, the greater the value the closer to the most recent common ancestor, i.e., origin of CRF47_BF. Note that these correspond well to the branch lengths observed leading to samples with <1000 days from diagnoses having longer branches from the root to the terminal samples and >3000 having shorter and more basal branches in the phylogram.

### 3.3 Analysis of drug resistance mutations

To identify drug resistance mutations in the CRF47_BF viruses, we analyzed the sequences with the Stanford HIV Drug Resistance Database’s HIVdb program (Tang et al., 2012). We found ARV drug resistance mutations in 5 patients: M184V or M184I mutations of resistance to nucleoside reverse transcriptase inhibitors (NRTI) in three samples; K103N mutation of resistance to nonnucleoside reverse transcriptase inhibitors (NNRTI) + K65N mutation of resistance to NRTIs in one sample; and E138A mutation associated with low level resistance to the NNRTI rilpivirine in one patient. Only one of these patients, with M184I mutation, was ARV drug-experienced.

### 3.4 Phylodynamics

With time-stamped sequence data, we performed a Bayesian Skygrid coalescent analysis to estimate historical population dynamics (Hill and Baele, 2019) of the CRF47_BF variants throughout Spain. Time labels (tipdates) were determined by the date of sample collection (ranging from 2007 to 2021). Our analysis supports a fairly dynamic population history of the CRF47_BF in Spain over the last 15 years with an initial increase in population size, a subsequent increase from 2011 to 2015, with a leveling off more recently, but seemingly increasing variance (Figure 4). This fluctuation in effective population size of CRF47_BF in Spain is consistent with the estimated incidence rates that also fluctuate considerably over this same time period (Figure 4), with an average effective population size estimated to be 155. Using BEAST, we estimated a chronogram to determine the time of origin for both the CRF47_BF clade as well as the timing of the introduction of CRF47_BF viruses to Spain (Figure 5). We estimated the origin of the CRF47_BF clade in Brazil (pP=1.0), dated to 1993-1994 (95% highest posterior density (HPD) interval between 1988 and 1998) and timed the introduction of CRF47 to Spain (pP=0.99) to be 1999-2000 (95% HPD interval between 1997-2002) (Figure 5). Similarly, viral strains seem to have entered once and spread through the Spanish regions of Basque Country (cluster I) (pP=1.0), Navarre (cluster II) (pP=1.0), and Aragon (cluster III) (pP=1.0) between 2008 and 2013 (Figure 5). These analyses, hence, suggest that CRF47_BF was probably circulating in Spain for over 10 years before it was identified through DNA sequencing, but clearly at a relatively low frequency. Given the sampling of CRF47 sequences, it appears that the introduction of this recombinant form to Spain was via Brazil supported by very high posterior probabilities (pP = 1.00).

**Figure 4.**
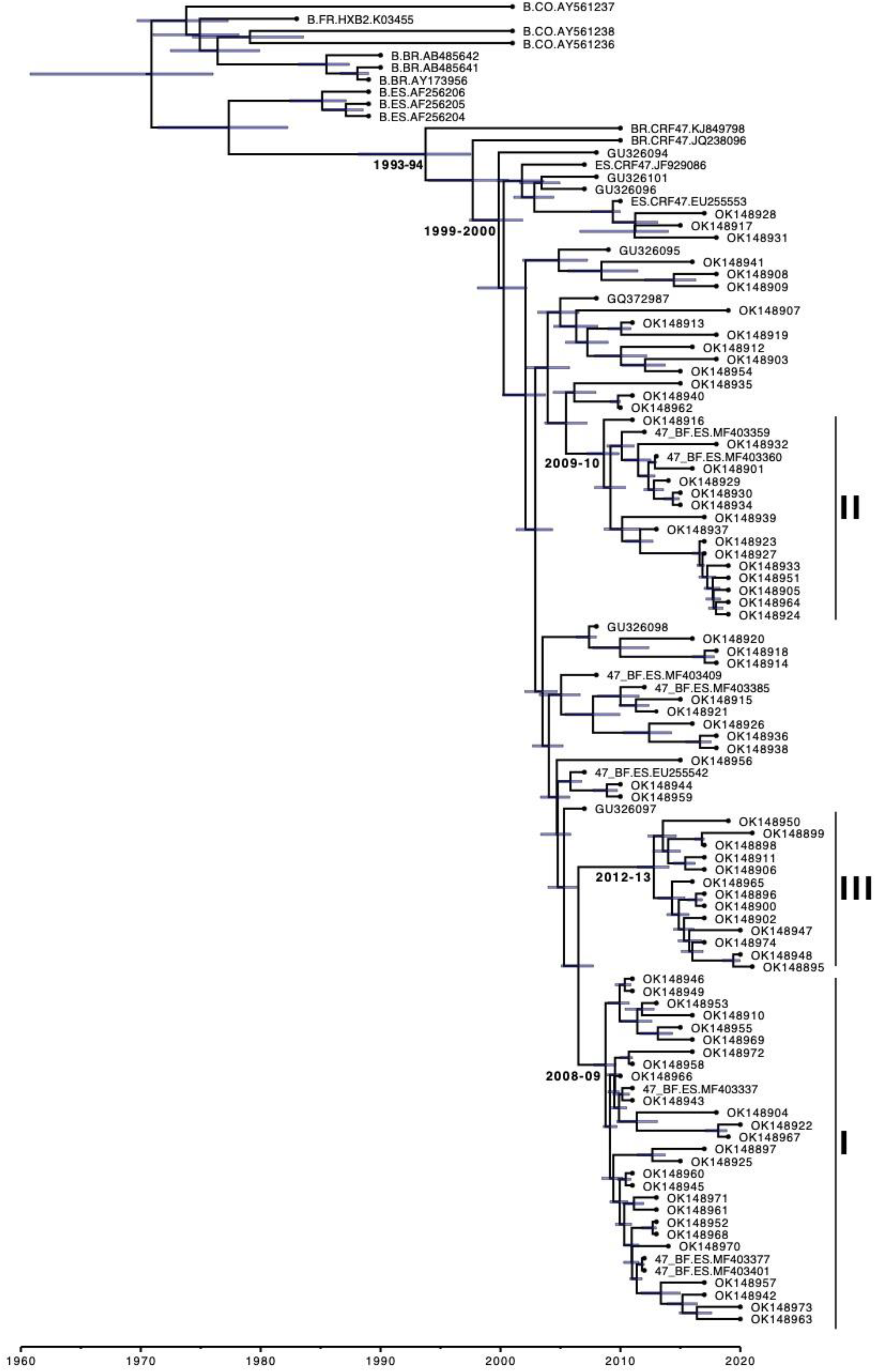
Population dynamics of CRF47_BF in Spain. Bayesian Skygrid estimate of fluctuating population size by year compared with the actual number of new CRF47_BF samples collected in the study each year.

## 4 Discussion

The HIV-1 CRF47_BF was first reported in 2010, detected in 9 samples collected in Spain in 2007-2009. Samples have subsequently been collected as this novel variant has spread throughout the country. Our phylogenetic analysis shows that isolates of CRF47_BF form a strongly supported monophyletic group (share a most recent common ancestor) relative to other CRFx_BF sequences, while all the CRFx_BF sequences form a weakly supported monophyletic group that also includes subtype B and subtype D, but, interestingly, not subsubtype F2. A focused phylogenetic analysis of the CRF47_BF sequences show a clear single origin in Brazil around 1993-94 with a subsequent transmission and rapid spread throughout Spain beginning around 1999-2000. Three strongly supported clusters, comprising a majority of viruses and associated with the regions of Basque Country, Navarre, and Aragon, were identified; this suggests that after a single introduction in Spain, CRF47_BF has spread mainly through localized point introductions and subsequent spread in different geographical areas. CRF47_BF is predominant in males (78%) with a predominantly heterosexual transmission (56% of the total, 61% of those with data on transmission mode). The phylodynamic analysis and epidemiological incidence data both support a fluctuating population size of CRF47_BF over the last 15 years with periods of expansion and contraction, suggesting that continued monitoring of this novel variant will be important to track its spread.

It is interesting to note that one cluster of 17 individuals, associated with Navarre, where 14 of 15 individuals with available data were male, was associated with transmission among MSM. Although three men were reported to be heterosexual, considering the great male preponderance in the cluster, it is probable that they are nondisclosed MSM (Hué et al., 2014; Ragonnet-Cronin et al., 2018). The identification of an MSM-associated cluster within the CRF47_BF clade may be indicative of the diffusion of CRF47_BF from a heterosexual-driven network to a MSM-driven network. A similar phenomenon has been observed for the two other CRFs of South American origin identified by us in Spain: CRF66_BF (Bacqué et al., 2021) and CRF89_BF (Delgado et al., 2021). Such phenomenon may reflect the migration of these CRFs from countries where heterosexual transmission is predominant to Spain, where most currently expanding HIV-1 clusters are associated with MSM (Patiño-Galindo et al., 2017; Gil et al., 2021^1^).

The recent expansion in Spain of CRF47_BF, whose Brazilian origin is first reported here, is one more example of the increasing relationship of the South American and European HIV-1 epidemics, also reflected in the propagation in Europe of other CRFs (12_BF, 17_BF, 60_BC, 66_BF, 89_BF) (Simonetti et al., 2014; Fabeni et al., 2015, 2020; Bacqué et al., 2021; Delgado et al., 2021) and variants of subtypes F1 and C (Tovanabutra et al., 2003; de Oliveira et al., 2010; Thomson et al., 2012; Lai et al., 2014; Carvalho et al., 2015; Delgado et al., 2015; Vinken et al., 2019) of South American ancestry, which probably derives from increasing migratory flows from South America to Europe.

The repeated introduction and expansion in Spain of multiple CRFs and non-B subtypes (Delgado et al., 2015; Delgado et al., 2019; Patiño-Galindo et al., 2017; Kostaki et al., 2019; González-Domenech et al., 2019) justifies the establishment of a HIV-1 molecular epidemiological surveillance system, aimed at promptly detecting the propagation of such variants, as well as rapidly expanding clusters, that could provide information in real-time on changes in the genetic composition and the dynamics of the HIV-1 epidemic to guide the implementation of preventive public health interventions (Paraskevis et al. 2016; German et al., 2017; Oster et al., 2018).

## Data Availability

All newly obtained sequences and associated data have been deposited in GenBank under accessions OK148895-OK148974

## 6 Author Contributions

MT, ED, MP-L conceived of the project. ED collected sequence data from the samples. GH, KC, MP-L, MT, ED conducted data analyses. HG performed data curation. SB, VM, MS, JC, and EG-B performed experimental work. The members of the Spanish Group for the Study of New HIV Diagnoses collected samples and clinical and epidemiological data for the study. KC, GH, MP-L wrote the original draft of the manuscript. MT, ED, HG edited the manuscript. All authors read and approved the manuscript.

## 8 Acknowledgments

We thank José Antonio Taboada, from Consellería de Sanidade, Government of Galicia, and Daniel Zulaika, from Unidad de Coordinación del Plan de Prevención y Control del SIDA, Osakidetza-Servicio Vasco de Salud, Government of the Basque Country, for their support of this study, and the personnel at the Genomic Unit, Instituto de Salud Carlos III, for technical assistance in sequencing.

## 9 Members of the Spanish Group for the Study of New HIV Diagnoses

**Hospital Universitario de Basurto**: Mª Carmen Nieto-Toboso; Silvia Hernáez*; Josefa Muñoz; Miren Zuriñe Zubero-Sulibarria; Sofía Ibarra-Ugarte; José Luis Díaz de Tuesta del Arco. **Hospital Universitario de Cruces, Bilbao**: Luis Elorduy; Leyre López-Soria; Elena Bereciartua-Bastarrica; Ane Josune Goikoetxea-Agirre. **Hospital Universitario de Galdakao**: Mª José López de Goikoetxea. **Hospital Universitario Donostia, San Sebastián**: Gustavo Cilla; José Antonio Iribarren; Yolanda Salicio; Maitane Aranzamendi. **Hospital Universitario Araba, Vitoria**: Carmen Gómez; José Joaquín Portu. **Hospital Universitario de Navarra, Pamplona**: Carmen Ezpeleta; Carmen Martín-Salas; Mª Gracia Ruiz-Alda; Aitziber Aguinaga. **Hospital Reina Sofía, Tudela**: José Javier García-Irure. **Hospital Universitario Sant Joan d’Alacant:** Fernando Buñuel; Francisco Jover-Díaz. **Complejo Hospitalario Universitario de Vigo**: Antonio Ocampo; Celia Miralles; Jorge Julio Cabrera. **Complejo Hospitalario Universitario de Pontevedra**: Matilde Trigo; Julio Diz-Aren. **Complejo Hospitalario Lucus Augusti, Lugo**: Ramón Rabuñal; Mª José Gude; Eva María Romay. **Complejo Hospitalario Universitario de Ourense**: Ricardo Fernández-Rodríguez; Juan García-Costa. **Hospital Universitario Miguel Servet, Zaragoza**: Ana María Martínez-Sapiña; Piedad Arazo. **Centro Sanitario Sandoval, Madrid**: Jorge del Romero. **Hospital Universitario Río Hortega, Valladolid:** Belén Lorenzo-Vidal. **Hospital Universitario de Toledo**: César Gómez.

*Current affiliation: Hospital Universitario Araba, Vitoria.

## 11 Data Availability Statement

The new sequence data generated as part of this study are available on GenBank via accession numbers OK148895-OK148974.

## Figure Legends

Figure 5. Bayesian (BEAST) chronogram estimate of CRF47_BF sequences from Spain and other CRF47_BF sequences from GenBank, as well as a few additional Subtype B sequences from Spain, Brazil, and Colombia plus HXB2, all to serve as an outgroup to the CRF47_BF sequences. Clusters associated with the Spanish regions of Basque Country (cluster I), Navarre (cluster II), and Aragon (cluster III) are indicated. Estimated years of emergence of CRF47_BF in Brazil, of its introduction in Spain, and of emergence of the Spanish clusters are indicated besides the corresponding nodes. 95% highest posterior density (HPD) intervals (blue bars) are shown for all time estimates.

## Footnotes

1 https://www.medrxiv.org/content/10.1101/2021.09.28.21264185v1

